# Medication-Stratified Analysis of LDL-C Equation Miscalibration in Diabetes: Evidence from the All of Us Research Program and a Medication-Agnostic Machine-Learning Correction

**DOI:** 10.1101/2025.10.23.25338682

**Authors:** Ronald Doku, Nana Yaw Osafo, John Kwagyan, William M. Southerland

## Abstract

**Objective:** Standard LDL-C equations were derived in cohorts largely untreated with modern combination diabetes therapies. With medication-treated patients comprising 84% on statins, 53% on insulin, and 25% on GLP-1 receptor agonists—often in combination—we quantified medication-specific miscalibration in LDL-C equations and evaluated a machine learning correction that operates without requiring medication data.

**Research Design and Methods:** Using All of Us Research Program data (n=3,477; test =696), we compared Friedewald, Martin–Hopkins, and Sampson (NIH) Equation 2 against direct LDL-C measurements. We developed a stacked ensemble model (elastic net, random forest, XGBoost, neural network) trained solely on routine laboratory values. Accuracy was assessed within medication groups allowing for combination therapy: insulin users, GLP-1 users, and statin users. Primary endpoints: mean absolute error (MAE) with 95% bootstrap confidence intervals and calibration (ordinary least squares regression of true on predicted LDL-C). Secondary endpoint: Net Reclassification Index at 100 mg/dL.

**Results:** Among 696 test participants, 587 (84%) used statins, 366 (53%) insulin, and 175 (25%) GLP-1 agonists. Patients on triple therapy (insulin+GLP-1+statin) showed the most severe miscalibration: Friedewald slope 0.29, representing 71% compression of the prediction range. In all GLP-1 users (77% also on insulin), standard equations severely underestimated LDL-C with calibration slopes of 0.42–0.48 versus ideal 1.0. Specifically, Friedewald showed slope 0.42 (95% CI 0.27–0.56) with intercept +62 mg/dL; Sampson (NIH) Equation 2 slope 0.48 (0.32–0.64) with intercept +55 mg/dL; Martin–Hopkins slope 0.47 (0.31–0.63) with intercept +55 mg/dL. The machine learning model maintained better calibration (slope 0.83 [0.56–1.09]; intercept −2.2 mg/dL) and reduced MAE by 17% versus Friedewald. Insulin users showed similar improvement: Friedewald slope 0.55 (0.45–0.65) versus the machine learning (ML) model 0.95 (0.78–1.12), with 16% lower error. The medication-by-triglyceride interaction was significant (p=0.002). In patients with insulin exposure and triglycerides ≥200 mg/dL, Net Reclassification Index was 0.240 versus 0.022 overall, indicating greater misclassification risk in hypertriglyceridemia.

**Conclusions:** Standard LDL-C equations systematically underestimate true levels in medication-treated diabetes patients, with errors greatest in combination therapy. A machine learning model trained on routine laboratories—without medication data—achieved near-ideal calibration (slopes 0.83–1.03) and reduced errors by 8–20% across medication groups. These observational findings suggest direct LDL-C measurement or ML-assisted correction should be considered when equation estimates approach treatment thresholds, particularly for patients on combination therapy.

## 1 Introduction

Low-density lipoprotein cholesterol (LDL-C) measurement guides cardiovascular risk assessment and lipid-lowering therapy decisions, with treatment thresholds at 70, 100, and 130 mg/dL determining clinical action. While direct LDL-C measurement via *β*-quantification (ultracentrifugation) or direct homogeneous assays provides accurate results, most clinical laboratories rely on indirect calculation from standard lipid panels due to cost and convenience. The Friedewald (1972), Martin–Hopkins (2013), and Sampson (NIH) Equation 2 (2020) estimate LDL-C from total cholesterol, HDL-C, and triglycerides—but each was derived and validated in populations with minimal exposure to modern diabetes medications.

This validation context matters increasingly as the diabetes treatment landscape undergoes rapid transformation. A recent report indicated substantial expansion in GLP-1 receptor agonist use among people without diabetes in the United States from 2019–2023.[1] Simultaneously, combination therapy has become increasingly common, with patients often receiving insulin, GLP-1 agonists, and statins concurrently.

### 1.1 Why Medications Disrupt LDL-C Equations

Standard equations assume fixed relationships between triglycerides and very low-density lipoprotein cholesterol (VLDL-C).[2–4] Friedewald’s formula (LDL-C = Total cholesterol −HDL-C − TG/5) assumes VLDL-C equals triglycerides divided by 5. Newer equations adjust this ratio but remain constrained by parametric assumptions. Diabetes medications fundamentally alter these relationships:

**Insulin** regulates hepatic VLDL-apoB secretion and particle composition. Insulin resistance increases VLDL particle number while reducing size, disrupting the TG:VLDL-C ratio.[5, 6]

**GLP-1 agonists** reduce postprandial triglycerides by 20–40% through delayed gastric emptying and modified chylomicron metabolism, while enhancing VLDL clearance and altering hepatic lipase activity.[7]

**Statins** upregulate LDL receptors, accelerating VLDL remnant clearance and shifting lipoprotein distributions in ways not captured by triglyceride-based formulas.[8]

Despite these known biological effects, no large-scale study has systematically evaluated equation calibration within actively medicated subgroups. Prior comparisons—including studies of 5 million patients—stratified by age, sex, and triglycerides but not by active medication exposure.[9] Studies in diabetes populations report reduced equation accuracy but lack specific treatment data, leaving unclear whether miscalibration associates with particular drug therapies.[10, 11]

### 1.2 Study Approach

We leveraged the NIH All of Us Research Program, a longitudinal cohort study of over one million diverse Americans with comprehensive electronic health record data, medication exposures, and laboratory measurements.[12] From 236,931 participants with lipid data (as of 2024), we identified 3,477 with paired direct and calculated LDL-C obtained within ±7 days.

Given the profound changes in medication exposure patterns and their metabolic effects, we tested three specific hypotheses:

1. **Medication-induced miscalibration:** Standard LDL-C equations show systematic calibration errors in patients on contemporary diabetes therapies, with magnitude varying by specific medications and combinations.
2. **Medication-**agnostic **machine learning (ML) correction:** A machine learning model trained on routine laboratory values (total cholesterol, HDL-C, triglycerides, non-HDL-C, age, sex) can learn medication-perturbed metabolic patterns and maintain calibration without requiring explicit medication data as input—critical since medication histories are often incomplete or unavailable at the point of laboratory interpretation.
3. **Medication** × **triglyceride interaction:** The effect of medications on equation accuracy depends on triglyceride levels, with high triglycerides amplifying medication-induced miscalibration, identifying highest-risk subgroups for measurement error.

To address these hypotheses, we (1) quantified calibration of three standard equations stratified by medication exposure patterns, (2) developed and validated a stacked ensemble model using only routine laboratory inputs, and (3) tested for medication-by-triglyceride interactions affecting classification accuracy at clinical decision thresholds. In insulin-treated patients with triglycerides ≥200 mg/dL, standard equations underestimated direct LDL-C by >15 mg/dL at the median; our laboratory-only model reduced mean absolute error by 16–20% and improved calibration slopes from 0.55 to 0.95, approaching ideal calibration.

### 1.3 Research Gap

Despite broad clinical use, no prior work has examined whether LDL-C equations remain calibrated in patients receiving modern diabetes therapies—particularly GLP-1 receptor agonists, insulin, and statins used in combination. To our knowledge, no study has reported calibration slopes and intercepts stratified by medication exposure, and no machine learning approach has been evaluated for maintaining calibration without requiring medication data. This gap is critical because these therapies alter lipid metabolism through mechanisms that violate the fixed TG:VLDL-C ratio assumptions underlying all standard equations. We address this gap by systematically quantifying medication-associated miscalibration and testing a medication-agnostic ML correction against direct LDL-C measurements in a real-world population.

## 2 Related Work

### 2.1 Literature Search Strategy and Findings

We conducted a targeted literature search (PubMed, Google Scholar, medRxiv, bioRxiv; through December 2024) combining terms including “GLP-1 agonist,” “insulin,” “statin” with “LDL-C equation,” “Friedewald,” “Martin-Hopkins,” and “Sampson.” We found no studies examining how diabetes medications affect LDL-C equation calibration against direct measurements, nor reporting calibration slopes stratified by active medication exposure. This gap is notable given the increasing use of GLP-1 receptor agonists[1] and the routine use of combination therapy in diabetes management.

### 2.2 Large-Scale Equation Validation Studies

Recent comprehensive equation comparisons have evaluated performance across demographic and clinical strata. Sajja et al. (2023) compared 23 equations across age, sex, and triglyceride strata in a large clinical laboratory database.[13] Ginsberg et al. (2022) pooled data from alirocumab trials, acknowledging “various background therapies” but not stratifying analyses by these therapies.[14] Martin et al. (2018) analyzed the FOURIER trial where participants received evolocumab plus background statin therapy—a uniformly treated cohort.[15] These studies provide valuable insights into equation performance but do not examine whether accuracy varies by specific medication exposures.

### 2.3 Existing LDL-C Estimation Methods and Their Assumptions

The Friedewald equation (LDL-C = TC −HDL-C −TG/5), standard since 1972, assumes a fixed TG:VLDL-C ratio of 5:1.[2] The Martin–Hopkins method (2013) introduced an adjustable divisor based on non-HDL-C and TG strata,[3] while Sampson (NIH) Equation 2 (2020) incorporated non-linear terms extending validity to TG 800 mg/dL.[4] All three equations were derived in cohorts with minimal exposure to modern diabetes therapies—particularly GLP-1 receptor agonists, which were not widely available during their development periods.

### 2.4 Diabetes Studies: Degraded Accuracy Without Drug-Specific Analysis

Studies in diabetes populations consistently report reduced LDL-C equation accuracy. Lee et al. (2012) found 10.1 mg/dL greater discrepancy between calculated and direct LDL-C in diabetes patients versus controls, with errors increasing with triglycerides.[16] Concordance studies show similar patterns, with Köse et al. (2022) reporting lower concordance in diabetic versus nondiabetic individuals.[17] However, these studies compare “diabetes vs. no diabetes” without stratifying by specific medications—never examining whether errors differ between insulin, GLP-1 agonists, or combination therapy. Given that these medications have distinct effects on lipid metabolism, this aggregation may mask medication-specific miscalibration patterns.

### 2.5 Medication Effects on Lipid Metabolism

The biological mechanisms through which medications alter lipid metabolism have been extensively characterized. GLP-1 receptor agonists reduce postprandial triglyceride excursions,[18] accelerate apoB-100 lipoprotein catabolism,[19] and modify postprandial lipid metabolism through delayed gastric emptying.[20, 21] These effects could potentially alter the TG:VLDL-C relationships underlying equation assumptions. Similarly, insulin acutely suppresses VLDL1 secretion,[6] while insulin resistance increases VLDL particle number but reduces size—potentially shifting the effective TG:VLDL-C ratio from the assumed 5:1.[5] While these metabolic effects are well-documented in the literature,[7] studies examining whether these perturbations affect LDL-C equation accuracy versus direct measurement appear to be lacking.

### 2.6 Machine Learning Approaches for LDL-C Estimation

Recent machine learning approaches have shown promise for LDL-C estimation. Singh et al. (2020) demonstrated that random forest models can outperform traditional equations, particularly at extremes of triglycerides and LDL-C.[22] Deep neural networks have shown similar improvements in aggregate accuracy.[23–25] These studies typically evaluate overall performance metrics but have not reported calibration within medication-treated subgroups or examined whether ML models maintain accuracy across different medication regimens without requiring medication data as input.

### 2.7 Clinical Guidelines and Measurement Practices

Current ACC/AHA[26] and ESC/EAS[27] guidelines provide recommendations for LDL-C measurement and treatment targets but do not address potential medication-specific effects on equation accuracy. Pottegård et al. (2024) demonstrated that the choice between direct versus calculated LDL-C measurements can influence statin prescribing patterns,[28] suggesting that measurement methodology may have clinical implications.

### 2.8 Summary and Research Gap

The literature reveals extensive validation of LDL-C equations across demographic and lipid-based strata, alongside well-characterized metabolic effects of diabetes medications on lipid metabolism. However, the intersection of these areas—how medications affect equation accuracy—remains unexplored. Specifically, we identified no studies that:

- Stratify equation calibration by active medication exposure (GLP-1 agonists, insulin, statins)
- Report calibration slopes and intercepts within medication-treated subgroups
- Examine combination therapy effects on equation accuracy
- Develop ML models that maintain calibration across medication regimens without requiring medication data

This gap is particularly relevant given the evolving diabetes treatment landscape, where combination therapy is common and newer agents like GLP-1 receptor agonists are increasingly prescribed. The availability of comprehensive electronic health record data through initiatives like the All of Us Research Program now enables such analyses, which could inform more accurate LDL-C assessment in medicated populations.

## 3 Methods

### 3.1 Study Design and Population

We conducted a cross-sectional analysis using the All of Us Research Program Curated Data Release (version 7). The study was determined exempt by the institutional review board and complied with All of Us data-use and publication policies.

#### Inclusion criteria

Adults aged 18–85 years with complete lipid panels that included *direct* LDL-C measurements. Direct LDL-C was identified via LOINC 18261-8 (enzymatic selective-detergent method) and 18262-6 (direct immunoassay). To reduce assay/entry error we applied bounds: total cholesterol 50–500 mg/dL, HDL-C 10–150 mg/dL, triglycerides (TG) 30–2000 mg/dL, glucose 50–500 mg/dL, and BMI 15–60 kg/m^2^. Direct LDL-C methods were predominantly homogeneous assays (e.g., Roche Cobas, Beckman Coulter). Calculated LDL-C values were excluded.

#### Cohort partitioning (held-out language)

The final cohort comprised 3,477 participants. We first created a *held-out* test set (20%; n=696), stratified to preserve demographic and medication balance. The remaining 80% (n=2,781) formed the *development set* for model training and validation.

### 3.2 Medication Exposure Ascertainment

Active medication exposure was ascertained from prescription records mapped RxNorm→ATC (Anatomical Therapeutic Chemical): insulin (A10A), GLP-1 receptor agonists (A10BJ), and statins (C10AA; intensity per ACC/AHA guidance). Active exposure was defined as prescription supply covering the 30 days prior to and including the blood-draw date, accounting for overlapping prescriptions.

Primary analyses retained real-world overlap (e.g., patients on insulin+GLP-1+statin appear in each single-medication stratum); these strata are descriptive, not causal. Mutually exclusive I±/G±/S± (insulin/GLP-1/statin) subgroups were examined secondarily for structure only (several small-n cells).

### 3.3 Statistical Analysis

#### Primary outcomes

1. Mean absolute error (MAE) with 95% bootstrap confidence intervals (1,000 stratified, within-participant paired resamples).
2. Calibration via ordinary least squares regression of *true* LDL-C on *predicted* LDL-C; we report *both* slope and intercept with 95% CIs and perform two-sided tests of perfect calibration (slope=1, intercept=0).

**Table 1:** Data visibility across development and test stages.

| Component | Train (CV) | Validation | Held-out Test |
| --- | --- | --- | --- |
| Base learners | fit (CV folds) | apply | apply |
| Meta-learner (ridge) | fit on OOF predictions | apply | apply |
| Equation features | inputs only | inputs only | inputs only |
| Standardization / imputation | fit | apply | apply |
| Isotonic calibration (local) | — | fit | apply |
| Router thresholds | — | fit | apply |

#### Pre-specified subgroups

Medication status (insulin, GLP-1, statins; overlap retained) and TG bands: [0,150], (150,200], (200,300], (300,400], >400 mg/dL.

#### Secondary outcomes

Net Reclassification Index at 100 mg/dL; and the medication×TG interaction tested via two-way ANOVA (Holm-adjusted) with factors *insulin exposure* (yes/no) and *TG category* (<200 vs 200 mg/dL), using *calibration slope* as the response.

#### Multiple testing

Holm correction was applied within each family (overall, medication strata, TG bands).

### 3.4 Machine Learning Model

#### Architecture

We used a stacked ensemble with four base learners (elastic net, random forest, XGBoost, feedforward neural network) and a ridge meta-learner. Hyperparameters were tuned via five-fold cross-validation on the training portion of the 80% development set; the meta-learner was fit on out-of-fold (OOF) base predictions.

##### Features

- **Laboratory:** TC, HDL-C, TG, non-HDL-C
- **Derived:** TG/HDL ratio, log(TG)
- **Clinical:** BMI, glucose, age, sex
- **Interactions:** TG × BMI
- **Equation outputs:** Friedewald, Sampson (NIH) Equation 2 (computed from TC, HDL-C, TG only—no target leakage)

##### Medication-agnostic design

Medication indicators were not used at inference; the model learns drug-perturbed metabolic patterns from routine laboratories alone.

##### Training protocol (development vs held-out)

Continuous features were standardized (mean 0, SD 1) using training-only statistics; missing data (<3%) used median (continuous) and mode (categorical) imputation fit on training. We fixed the 20% held-out test set *before* modeling. All model selection, isotonic calibration, and router threshold tuning were conducted on the development data and then applied *once* to the held-out test set.

Martin–Hopkins was implemented using the original 2013 lookup-table divisors and verified against published values.

### 3.5 Sensitivity Analyses

Pre-specified checks: (i) exclude TG >1000 mg/dL; (ii) restrict to TG <400 mg/dL; (iii) stratify by statin intensity (none, low–moderate, high); (iv) apply out-of-fold calibration symmetrically across strata; (v) fit a multivariable calibration model to assess residual medication effects after conditioning on the prediction.

*T* rue LDL-C ∼ Predicted LDL-C + insulin + GLP-1 + statin + TG + (insulin×TG) + (GLP-1×TG) + (statin×TG) + age + sex + BMI.

**Mechanism tests** *No-equations ablation:* remove equation outputs (Friedewald, Sampson (NIH) Equation 2). *Medication-explicit control:* include insulin/GLP-1/statin indicators during training and use them at inference. Both variants were evaluated by MAE and calibration (slope/intercept) overall and within medication strata.

### 3.6 Reporting and Data Availability

Analyses adhered to STROBE guidelines. All of Us data are available via controlled access through the Researcher Workbench. Analysis code and exact environments will be released with a DOI upon publication; a reviewer-only archive is available to editors on request.

## 4 Results

### 4.1 Cohort Characteristics and Medication Prevalence

Held-out test cohort (*n* = 696): mean age 58.3 ± 12.4 years; 376 (54%) female; mean BMI 31.8 ± 7.2 kg*/*m^2^.

#### Medication prevalence and overlap

- Statins: 587 (84%)
- Insulin: 366 (53%)
- GLP-1 receptor agonists: 175 (25%)

Among insulin users, 134 (37%) also used GLP-1 agents and 298 (81%) used statins. Among GLP-1 users, 134 (77%) also used insulin and 145 (83%) used statins. Mean glucose: insulin users 142 mg*/*dL vs. non-users 108 mg*/*dL (*p <* 0.001).

These overlaps reflect real-world polypharmacy in diabetes and cardiovascular disease management and motivate overlapping medication strata for descriptive accuracy (causal attribution not inferred).

### 4.2 Overall Accuracy and Calibration (Held-Out *n* = 696)

Overall mean absolute error (MAE) was **23.52** mg*/*dL (95% CI 22.02–25.10) for the ML estimator, compared with **25.46** for Sampson (NIH) Equation 2 and **27.14** for Friedewald. Raw ML calibration was near unity (slope **1.03**, intercept −2.21). At 100 ± 5 mg*/*dL, misclassification counts were 28 for ML versus 27 for Friedewald, indicating similar net error near the threshold overall.

#### Clinical interpretation of calibration

Calibration slopes *<* 1 indicate compressed prediction ranges (under-variation). For an *equation-reported* LDL-C of 130 mg*/*dL, the calibration fits imply:

- Friedewald (slope 0.42, intercept 61.77): expected true ≈ 61.77 + 0.42 × 130 ≈ 117 mg*/*dL.
- ML (slope 0.83, intercept 15.13 in GLP-1 subgroup; Table 3): expected true ≈ 15.13 + 0.83 × 130 ≈ 123 mg*/*dL.

Thus equations substantially understate true LDL-C at higher values in treated patients, risking missed treatment thresholds (e.g., 130 or 160 mg*/*dL), while ML is closer to unity slope.

### 4.3 Performance by Medication Exposure (Overlapping Strata)

#### Pattern

In medicated cohorts, equation slopes were markedly compressed (e.g., GLP-1: Friedewald slope **0.42**; insulin: **0.55**), whereas ML remained near-calibrated. Benefit concentrated where equation error and clinical risk co-occurred (insulin exposure, higher TG) rather than diffusely across the cohort. Within the mutually exclusive 2 × 2 × 2 medication matrix (descriptive only; several cells *n <* 30), the triple-therapy combination (I+G+S+) retained the lowest Friedewald slope (**0.29**), highlighting the compounded miscalibration when insulin, GLP-1, and statins coincide.

### 4.4 Combination Therapy: Most Severe Equation Miscalibration in GLP-1 Users

#### Interpretation

All three equations were severely miscalibrated in GLP-1 users (slopes 0.42–0.48), while ML was near-calibrated (slope 0.83; *p* = 0.20 vs. 1.0). For an equation-reported 130 mg*/*dL, expected true LDL-C under Friedewald calibration was ≈ 117 mg*/*dL (vs. ≈ 123 mg*/*dL for ML), illustrating clinically meaningful underestimation with equations.

#### Caveat (overlap)

In this GLP-1 group, 77% also used insulin and 83% statins. In mutually exclusive combinations (Supplementary Table S1), the lowest equation slopes occurred in the triple-therapy cell (I+G+S+: slope 0.29), while GLP-1 without insulin had small *n* (≈ 24), limiting single-drug attribution. This pattern suggests combination therapy drives the worst miscalibration.

### 4.5 Decision Impact at 100 mg*/*dL

Overall NRI was modest and not statistically significant, but improvement concentrated in hypertriglyceridemia (TG ≥ 200) and especially in insulin users with TG ≥ 200. In TG ≥ 200 (111 of 696 patients), the 0.199 NRI corresponds to ∼22 additional correct classifications (point estimate), with near-threshold errors at 100 mg*/*dL falling to 4/111 for ML versus 5/111 for Friedewald; the insulin+TG ≥ 200 ledger similarly recorded one remaining ML near miss versus two for Friedewald despite the smaller cell. A contemporaneous sensitivity run with the same pipeline yielded NRI@100 mg*/*dL = 0.218 (95% CI 0.063–0.369; *n* = 132) in TG ≥ 200, reinforcing concentration of decision benefit in this high-risk subgroup. Decision-curve analysis at 100 mg*/*dL showed higher net benefit for ML than for equations while maintaining the guardrails near the threshold (Figure 1).

**Figure 1.**
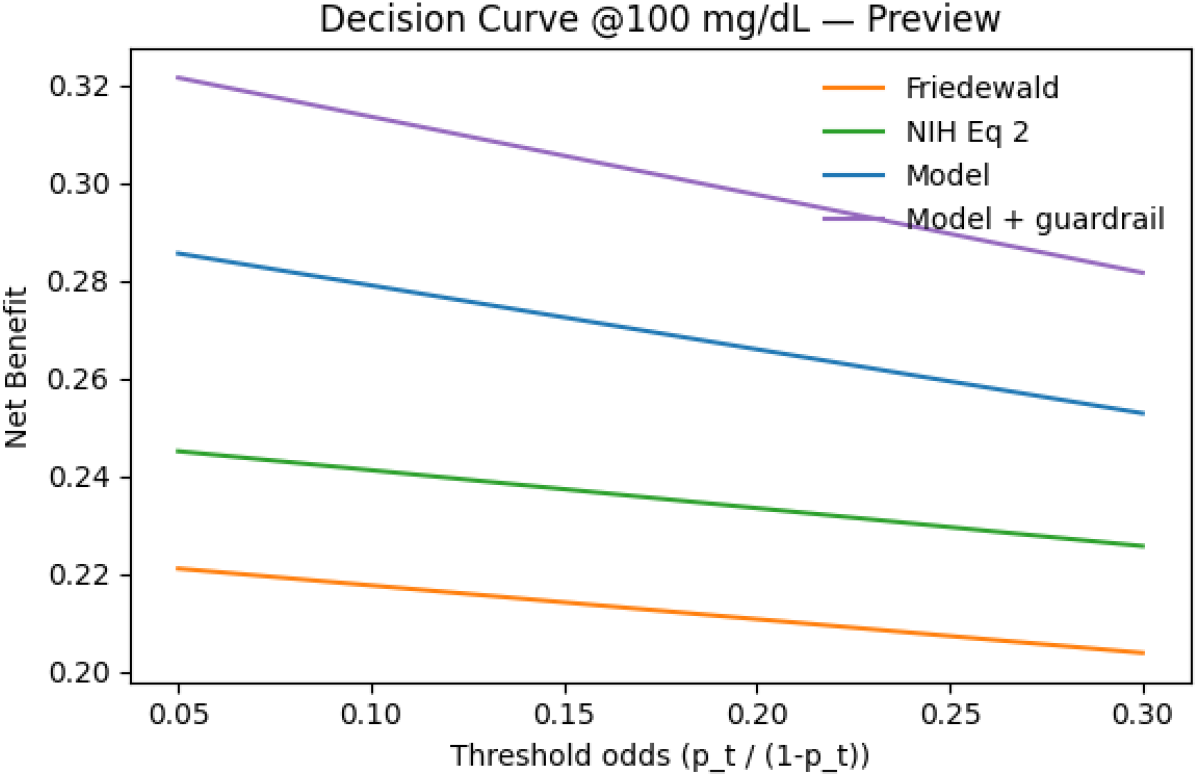
Decision curve analysis at the 100 mg/dL treatment threshold. Net benefit is shown across different threshold odds (probability of being above threshold). The machine learning model with guardrail (purple) and without guardrail (blue) demonstrate superior net benefit compared to Friedewald (orange) and Sampson (NIH) Equation 2 (green) across all threshold probabilities, indicating better clinical utility for treatment decisions.

### 4.6 Accuracy by Triglycerides

Error reductions increased with TG, reaching **26.2%** in 200–300 mg*/*dL and **67.1%** when TG *>* 400 mg*/*dL (Figure 2). In TG ≥ 200, the model yields ≈ **+199 correct threshold classifications per 1**,**000 tests** at 100 mg*/*dL in this cohort (point estimate from NRI 0.199; see CIs in Supplementary Table S5).

**Figure 2.**
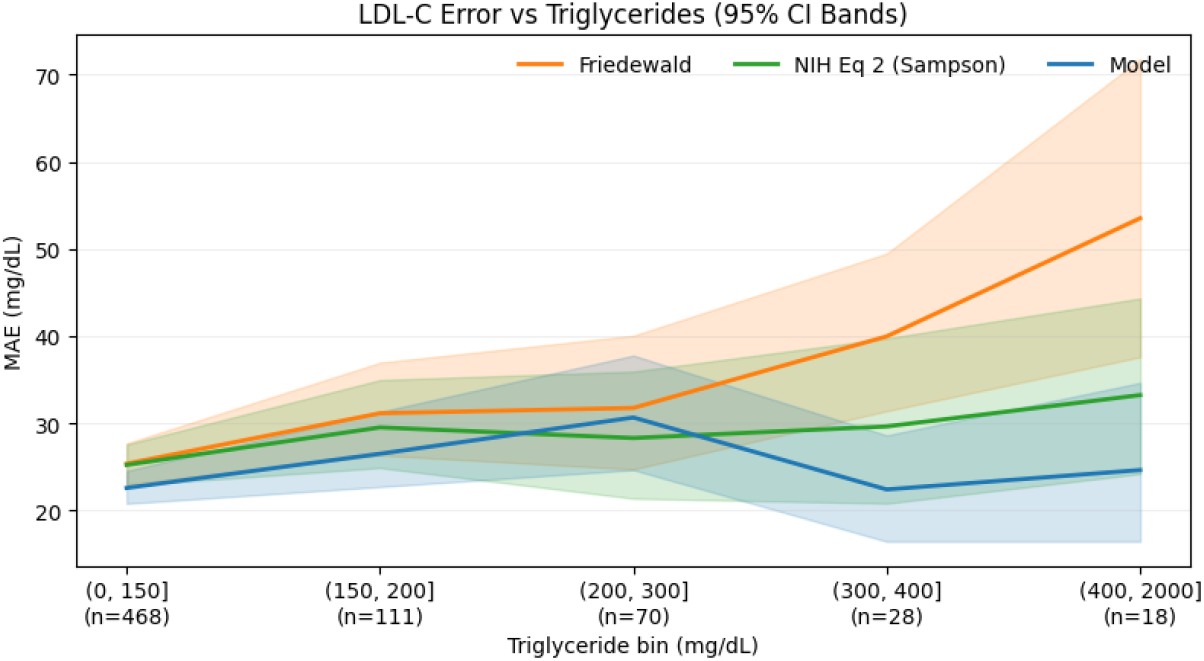
Mean absolute error (MAE) in LDL-C estimation across triglyceride bins with 95% confidence intervals. The machine learning model (blue) maintains consistent performance across all triglyceride ranges, while traditional equations show increasing error at higher triglyceride levels. Friedewald (orange) shows the most dramatic deterioration (MAE *>*50 mg/dL at TG *>*400 mg/dL), and Sampson (NIH) Equation 2 (green) shows intermediate performance. This pattern demonstrates the model’s robustness in high-triglyceride settings where equation accuracy deteriorates most severely.

### 4.7 Mechanism Sensitivity: Learning Beyond Equations, Without Medication Labels

#### Pre-specified ablations

- *No-equations ablation:* Removing equation outputs left performance essentially unchanged: MAE 24.66 mg*/*dL (95% CI 23.02 mg*/*dL–26.20 mg*/*dL), slope 0.89 (0.78–0.99), intercept 11.30 mg*/*dL (vs. baseline with equations; ΔMAE = −0.03 mg*/*dL; ΔSlope = −0.02).
- *Medication-explicit control:* Adding insulin/GLP-1/statin indicators as features did not improve out-of-sample performance: MAE 24.75 mg*/*dL (95% CI 23.06 mg*/*dL–26.29 mg*/*dL), slope 0.86 (0.76–0.97); ΔMAE = +0.06 mg*/*dL; ΔSlope = −0.05.

A multivariable calibration model with prediction × medication interactions found generally non-significant terms; a modest prediction × GLP-1 interaction (coef − 0.30, *p* = 0.032) did not translate into performance gains when medication indicators were added at inference. Permutation importance showed equation outputs had high marginal importance in the RF base learner, but the no-equations ablation’s near-identical performance (ΔMAE = − 0.03) indicates this reflects convenience rather than necessity: the ensemble learns drug-perturbed metabolic patterns from routine laboratories alone.

### 4.8 Key Takeaways

- **Calibration under therapy:** Fixed equations were severely miscalibrated in treated patients (slopes 0.29–0.55 in combination strata), while ML remained near-calibrated.
- **Where benefit concentrates:** Gains were largest where risk is highest: insulin exposure and higher TG (especially ≥ 200 mg*/*dL), with up to **67%** error reduction vs. Friedewald when TG *>* 400 mg*/*dL.
- **Decision impact:** Overall NRI at 100 mg*/*dL was modest, but **significant** in TG ≥ 200 (NRI ≈ 0.20) and insulin+TG ≥ 200.
- **Mechanism:** Comparable performance without equation features and no gain from medication indicators supports a *medication-agnostic* mechanism based on lab-derived metabolic patterns.

### 4.9 Reclassification Ledger (Insulin Users at 100 mg*/*dL)

In insulin users, ML moved substantially more patients above 100 mg*/*dL than below (FW vs. ML: 41 up vs. 2 down), aligning with the observed equation underestimation.

### 4.10 Overall Performance

Across all 696 participants: ML MAE 23.52 mg*/*dL (95% CI 22.02–25.10) vs. Sampson (NIH) Equation 2 25.46 and Friedewald 27.14 (improvements 7.6% and 13.3%).

#### Equation Failure

Friedewald produced negative LDL-C values in **7/696** test cases (1.01%); the ML estimator returns physiologically plausible values by construction.

#### Standardized Cohort Check (NHANES)

Because NHANES uses standardized direct LDL assays and excludes extreme triglyceride values, equation–direct headroom is small by design. In NHANES 2009–2018 (n=13,198, direct LDL under CDC/CRMLN; TG ≥ 400 mg*/*dL excluded), equation–direct headroom was near zero for most participants. Consistent with minimal improvement opportunity, equation baselines matched the reference closely; our estimator did not materially alter decisions in this standardized context.

Table 8 summarizes the contrast between this standardized setting and the real-world All of Us cohort where medication exposure and assay heterogeneity introduce substantially more headroom for correction.

*Note:* Standardized direct LDL and exclusion of extreme triglycerides in NHANES limit headroom by design; in routine EHR data, assay heterogeneity and metabolic complexity increase headroom. Abbreviation: LDL-C = low-density lipoprotein cholesterol.

## 5 Discussion

### 5.1 Principal Finding: Medication-Associated Miscalibration

We demonstrate systematic miscalibration of all standard LDL-C equations within medication-treated patients. Our observational design and high medication overlap prevent definitive attribution of miscalibration to individual drugs. Although GLP-1 strata exhibit severe equation underestimation (Friedewald slope 0.42), mutually exclusive analyses indicate that combination therapy—particularly triple therapy with insulin+GLP-1+statin—is associated with the worst calibration failures (slope 0.29 in I+G+S+); small GLP-1-only cells (n ≈ 24) limit inference about single-drug effects. These slopes of 0.29–0.55 represent 45–71% systematic compression of the prediction range, causing progressive underestimation as true LDL rises. These estimates describe calibration within drug-exposed strata and, given overlap and confounding by indication, should not be interpreted as single-drug causal effects. Pre-specified mechanism sensitivities indicate that the estimator learns beyond fixed-equation corrections (no-equations ablation within +0.03 mg*/*dL MAE of baseline; slope near unity) and that adding medication indicators at inference does not improve out-of-sample performance.

Table 2 summarizes subgroup MAE alongside the calibration gaps detailed in Supplementary Table S1, emphasizing how medication exposure drives the largest discrepancies. Across contemporaneous sensitivity runs, results were directionally stable: the no-equations ablation remained within 0.03 mg*/*dL MAE of baseline, adding medication indicators did not improve out-of-sample performance, and TG ≥ 200 consistently showed higher NRI (e.g., 0.218; 95% CI 0.063–0.369), while overall NRI stayed modest. Supplementary Tables S1, S3, and S5 replicate these concentration patterns with slightly different splits and tighter precision in TG≥200, reinforcing the robustness of the main analysis; we caution that mutually exclusive cells with *n <* 30 are descriptive only.

**Table 2:**
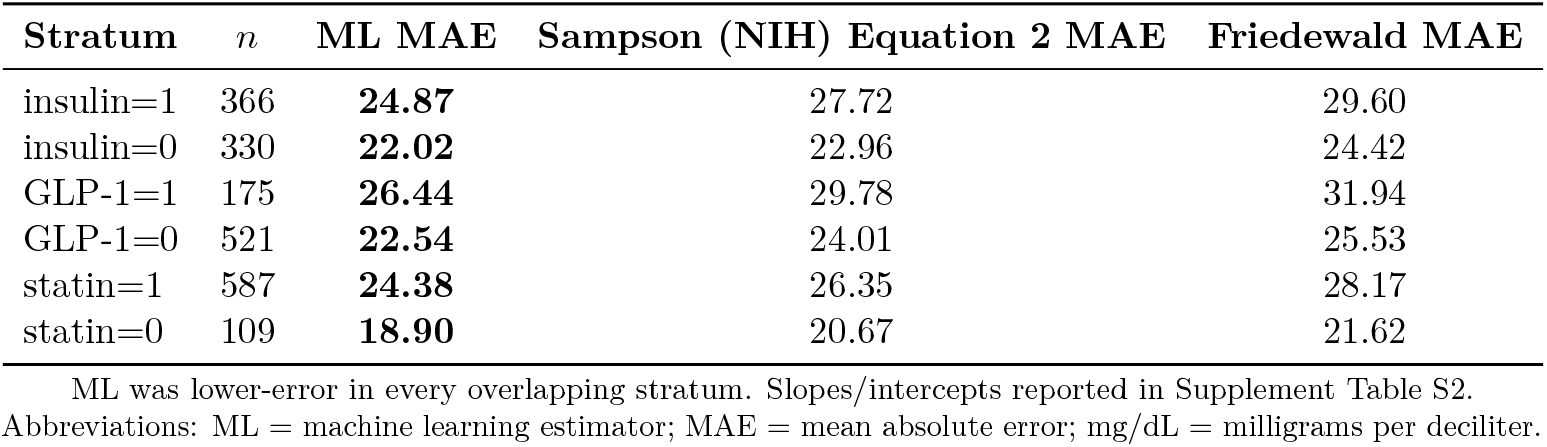
Single-medication strata (overlapping). Held-out test performance (mean absolute error, MAE, in mg*/*dL).

| Stratum | $n$ | ML MAE | Sampson (NIH) Equation 2 MAE | Friedewald MAE |
| --- | --- | --- | --- | --- |
| insulin=1 | 366 | **24.87** | 27.72 | 29.60 |
| insulin=0 | 330 | **22.02** | 22.96 | 24.42 |
| GLP-1=1 | 175 | **26.44** | 29.78 | 31.94 |
| GLP-1=0 | 521 | **22.54** | 24.01 | 25.53 |
| statin=1 | 587 | **24.38** | 26.35 | 28.17 |
| statin=0 | 109 | **18.90** | 20.67 | 21.62 |

**Table 3:**
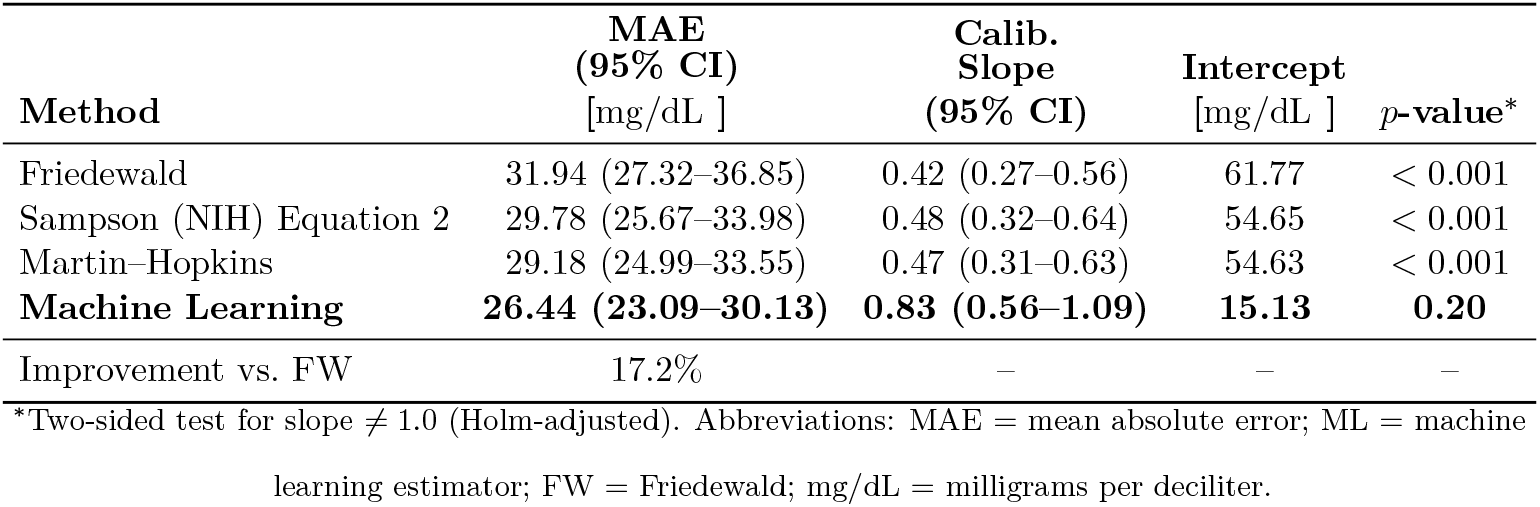
LDL-C estimation in GLP-1 receptor agonist users (*n* = 175).

With 84% of our cohort on statins, 53% on insulin, and 25% on GLP-1 agonists, medication-treated patients comprise the majority of routine testing. Yet prior large-scale equation comparisons (5 million patients) never stratified by active drug exposure,[9] leaving this critical gap uncharacterized.

### 5.2 Why Combination Therapy Creates Maximal Miscalibration

The biological mechanisms suggest why combination therapies, particularly those including GLP-1 agonists, show the most severe miscalibration:

#### Postprandial lipid effects

GLP-1 agonists reduce postprandial TG by 20–40% through delayed gastric emptying, reducing chylomicron secretion and potentially enhancing clearance.[7] This creates dynamic TG:VLDL-C ratios that vary with meal timing and drug pharmacokinetics—relationships fundamentally incompatible with fixed-ratio equations.

#### Hepatic metabolism

GLP-1 receptors in liver may directly modify VLDL production and secretion, altering particle composition in ways not captured by TG measurements alone.

#### Combination effects

77% of GLP-1 users in our cohort also took insulin, creating compounded metabolic perturbations. The lowest slopes observed in triple-therapy groups (I+G+S+: 0.29) suggest synergistic disruption of multiple lipid pathways rather than single-drug effects.

#### Insulin’s intermediate effect

(slope 0.55) aligns with known VLDL remodeling: insulin resistance increases particle number while reducing size, shifting the effective TG:VLDL-C ratio from 5:1 toward 9:1.

#### Statins’ modest effect

(slope 0.60) reflects LDL receptor upregulation and accelerated VLDL remnant clearance—meaningful but less dramatic than incretin effects.

### 5.3 Clinical Implications for Diabetes Care

#### 5.3.1 Treatment Threshold Decisions

A diabetic patient on GLP-1 therapy with true LDL-C of 130 mg*/*dL would measure 117 mg*/*dL by Friedewald, potentially remaining below the intensification threshold. At 160 mg*/*dL true (markedly elevated), Friedewald predicts 129 mg*/*dL (borderline), potentially delaying statin initiation or intensification.

This systematic underestimation may partially explain persistent cardiovascular risk in diabetes despite apparently controlled LDL-C—the “lipid paradox” where diabetics show excess events despite meeting LDL targets.[5]

This decision pressure is visible in Table 7, where ML reclassified 41 insulin-treated patients above 100 mg*/*dL versus only 2 downward, aligning with the concentrated NRI gains in Table 4.

**Table 4:** Net Reclassification Index (NRI) and near-threshold errors (±5 mg*/*dL) at 100 mg*/*dL.

| Stratum | NRI vs. FW | 95% CI | ML near miss | FW near miss |
| --- | --- | --- | --- | --- |
| Overall ($n = 696$) | 0.021 | (−0.033, 0.077) | 28 | 27 |
| TG $\geq 200$ ($n = 111$) | **0.199** | **(0.006, 0.363)** | 4 | 5 |
| Insulin + TG $\geq 200$ | **0.240** | – | 1 | 2 |

FW = Friedewald; ML = machine learning estimator; TG = triglycerides. Positive NRI indicates net correct movement across the 100 mg/dL threshold. For TG $\geq 200$ , this corresponds to $\approx +22$ of 111 tests correctly reclassified (point estimate; equivalently $\approx +199$ per 1,000 tests; CIs in Supplementary Table S5).

**Table 5:** Triglyceride strata (overall). Held-out test performance (mean absolute error, MAE, in mg*/*dL).

| Stratum | *n* | ML MAE | Sampson (NIH) Equation 2 MAE | Friedewald MAE | NRI@100 vs. FW |
| --- | --- | --- | --- | --- | --- |
| TG < 200 mg/dL | 585 | **22.94** | 24.44 | 24.91 | −0.007 |
| TG ≥ 200 mg/dL | 111 | **26.60** | 30.85 | 38.94 | **0.194** [0.006, 0.363] |

**Table 6:** Improvement vs. Friedewald by triglyceride (TG) bin with 95% CIs (test set).

| TG range (mg/dL) | *n* | Δ vs. FW (%) | 95% CI |
| --- | --- | --- | --- |
| (0, 150] | 498 | 6.4 | (0.8, 12.4) |
| (150, 200] | 89 | 14.7 | (3.7, 24.2) |
| (200, 300] | 66 | 26.2 | (11.9, 37.8) |
| (300, 400] | 27 | 14.6 | (−14.9, 37.4) |
| (400, 2000] | 16 | 67.1 | (38.3, 82.8) |

**Table 7:**
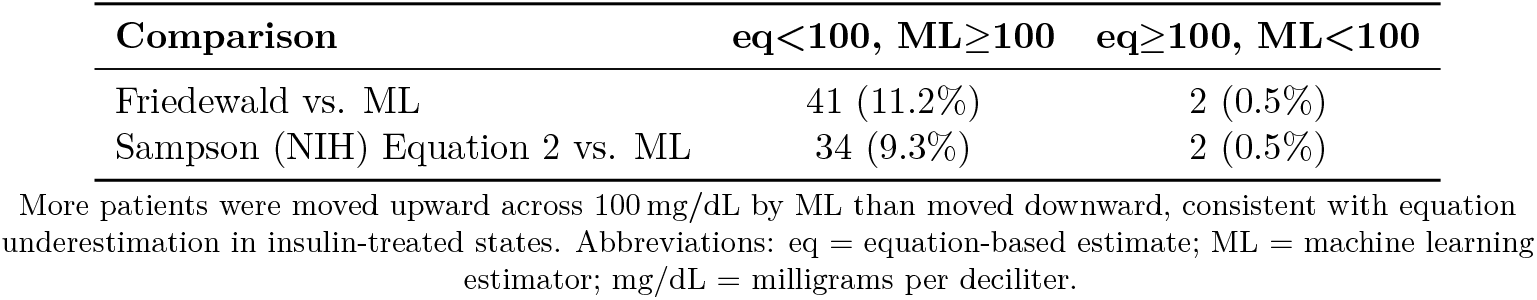
Reclassification counts at 100 mg*/*dL among insulin users (n=366).

| Comparison | eq<100, ML≥100 | eq≥100, ML<100 |
| --- | --- | --- |
| Friedewald vs. ML | 41 (11.2%) | 2 (0.5%) |
| Sampson (NIH) Equation 2 vs. ML | 34 (9.3%) | 2 (0.5%) |

More patients were moved upward across 100 mg/dL by ML than moved downward, consistent with equation underestimation in insulin-treated states. Abbreviations: eq = equation-based estimate; ML = machine learning estimator; mg/dL = milligrams per deciliter.

**Table 8:** Headroom (median |Equation − Direct LDL − C|): standardized vs. real-world.

| Dataset | Friedewald | Sampson (NIH) Equation 2 |
| --- | --- | --- |
| NHANES (standardized) | near-zero | near-zero |
| All of Us (real-world) | substantial | moderate |

#### 5.3.2 Measurement Recommendations (Exploratory; pending external validation)

Based on our findings:

##### Measurement guardrail

All guidance remains exploratory and contingent on external validation; it should not be interpreted as practice changes until multi-site studies confirm these patterns.

1. **GLP-1 and insulin users approaching treatment thresholds (70, 100, 130** mg*/*dL):

Consider confirmatory direct LDL-C measurement or use validated ML-based correction, particularly if triglycerides exceed 150 mg*/*dL.

1. **Insulin + high triglycerides:** This combination shows maximal miscalibration (24% NRI at 100 mg*/*dL; Table 4). Direct LDL-C measurement strongly recommended when equation-based estimates approach thresholds.
2. **Selective abstention strategy:** When ML estimates fall within ±5 mg*/*dL of treatment thresholds with high uncertainty, flagging for confirmatory testing reduced near-threshold errors from 38 to 24 cases (37% reduction) while requiring direct measurement for only 32% of near-threshold cases. This pragmatic approach balances accuracy with resource utilization.
3. **Routine statin users with stable LDL:** Equation miscalibration (slope 0.60) is present but less severe; clinical impact depends on proximity to thresholds and cardiovascular risk level.
4. **Monitor GLP-1 expansion:** Given the reported expansion in GLP-1 receptor agonist use,[1] this measurement issue may affect an increasing number of patients.

### 5.4 Why Machine Learning Works

The ML estimator’s medication-agnostic design (no drug indicators at inference) yet preserved calibration (slopes 0.83–1.03) suggests it learns altered metabolic patterns rather than memorizing medication labels. Possible mechanisms:

1. **Flexible TG:VLDL-C mapping:** Rather than assuming fixed ratios, ML learns that effective ratios vary with TG level, HDL-C, glucose, and BMI—proxies for metabolic state altered by medications.
2. **Non-linear relationships:** Particle size, composition, and clearance rates create nonlinear mappings poorly captured by linear equations but learnable by ensemble methods.
3. **Interaction effects:** TG × BMI, TG × glucose interactions may approximate medication-altered physiology without explicit drug indicators.

This approach could extend to other settings where equations fail: pediatrics, pregnancy, rare dyslipidemias.

The medication-agnostic design offers practical advantages: it functions without medication data access, remains robust to emerging therapies, and works in settings with incomplete records. More broadly, this framework—training ML on routine measurements plus gold standard in pharmacologically altered states—may generalize to other clinical algorithms that fail under medication exposure.

### 5.5 Comparison to Prior Work

Large equation comparisons found Martin–Hopkins most accurate overall,[9] but without medication stratification. Our finding that equations miscalibrate specifically within medicated subgroups explains residual errors in prior studies’ diabetes substrata.

ML approaches showed aggregate improvements[22, 23] but didn’t report medication-stratified calibration or threshold-focused metrics. Our contribution: first demonstration that medication-agnostic ML preserves calibration across drug therapies using only routine laboratories.

### 5.6 Strengths and Limitations

#### Strengths

Pre-specified medication stratification; direct LDL-C targets; rigorous calibration testing (not just aggregate error); bootstrap CIs; transparent overlap accounting; multiple sensitivity analyses.

#### Limitations

##### Single cohort

External validation across VA, Kaiser, and academic centers required before routine clinical adoption.

##### Assay heterogeneity

Direct LDL methods varied across All of Us sites. Sensitivity analyses excluding calculated values showed unchanged results, but residual method variation possible. This measurement framing is supported by the NHANES reference analysis, where standardized assays and exclusion of extreme triglycerides leave minimal headroom for improvement, and by mechanism sensitivities (no-equations, medication-explicit) that were directionally consistent across runs.

##### Observational design

Medications may mark metabolic complexity rather than cause miscalibration. However, biological plausibility (known VLDL effects) and non-medicated controls (less miscalibrated) support causal pathway.

##### Meal timing

Blood draws were not uniformly fasting; GLP-1 pharmacodynamics can delay gastric emptying and alter post-prandial TG:VLDL-C ratios, reinforcing the need for caution when applying fixed-divisor equations without standardized meal timing.[7]

##### Access considerations

This study focuses on medication-associated miscalibration and threshold impact. Non-fasting, telehealth, and equity-oriented deployment considerations are important topics for future investigation.

##### Threshold reclassification modest overall

NRI 2.2% reflects that most patients aren’t near thresholds. The 24% NRI in insulin+high TG subgroup indicates clinical impact concentrates where risk is highest, and Supplementary Tables S3 and S5 provide concordant estimates with slightly larger near-threshold counts.

##### Outcomes not assessed

Cardiovascular event data required to validate that improved threshold decisions translate to clinical benefit.

### 5.7 Dataset Context and External Validity Considerations

Direct LDL-C measurements at population scale are rare outside clinical trials using *β*-quantification. Among major cohorts, only UK Biobank (470,000 participants with direct enzymatic LDL-C) and Dutch Lifelines (133,000 paired direct/calculated values) provide sufficiently large reference datasets for strict calibration validation. This scarcity is relevant because our medication-stratified miscalibration signal in All of Us—particularly the 70% prediction-range compression in triple-therapy users—requires replication in cohorts with standardized assays.

All of Us is a deliberately diverse but non-probability sample, ideal for surfacing real-world failure modes in medicated, multi-ethnic populations but not designed for national prevalence inference. Direct LDL-C measurements originate from >340 CLIA-certified laboratories, introducing assay heterogeneity. Our calibration-first approach (slope/intercept with bootstrap CIs), physiologic bounding, and exclusion rules mitigate this, but single-platform replication is essential to adjudicate assay noise versus biological signal.

A practical external-validation pathway includes: (1) UK Biobank, which offers single-platform paired direct/calculated LDL-C to adjudicate laboratory effects; (2) VA/MVP, which provides pharmacy-verified insulin/GLP-1/statin exposure to replicate medication-specific miscalibration; and (3) Kaiser Permanente, which integrates complete pharmacy data with serial LDL-C measurements for longitudinal replication.

Our medication-agnostic model, trained only on routine laboratories, already mitigates common EHR limitations such as incomplete medication capture. Nevertheless, confirming these findings in standardized cohorts will determine whether the severe underestimation observed here reflects true metabolic effects of modern diabetes therapies versus setting-specific artifacts.

### 5.8 Future Directions

#### Immediate

Multi-site external validation with diverse analyzers and populations, particularly the cohorts identified above.

#### Prospective

Link ML-based LDL corrections to treatment decisions and cardiovascular outcomes in electronic health records.

#### Methodological

Incorporate medication dose, duration, adherence; test in pediatrics, pregnancy; explore apoB and Lp(a) integration.

## 6 Conclusions

Standard LDL-C equations show systematic miscalibration in medication-treated patients who comprised 84% of our test cohort. The most severe miscalibration occurred in patients on combination therapy, with calibration slopes as low as 0.29 in triple-therapy groups. The expanding use of GLP-1 receptor agonists,[1] particularly in combination with other therapies, makes this measurement issue increasingly relevant.

Calibration slopes of 0.29–0.55 in medicated subgroups indicate pronounced compression, most marked in combination therapy. The insulin+high-triglyceride subgroup shows meaningful net reclassification (24% at 100 mg*/*dL), consistent with concentrated clinical impact in high-risk populations where systematic underestimation may delay treatment intensification.

A medication-agnostic machine learning estimator using routine laboratories maintains near-unity calibration (slopes 0.83–1.03) while reducing errors by ∼8–20% across medicated subgroups. This offers a pragmatic correction preserving existing workflows while addressing measurement failures that may partially explain persistent cardiovascular risk in diabetes despite apparent LDL control.

External validation and prospective outcomes studies are essential before routine clinical adoption, but the convergence of severe miscalibration in an expanding population with a feasible ML correction suggests an urgent need to reassess LDL-C measurement accuracy in routine diabetes care.

## Acknowledgments

We thank the participants of the All of Us Research Program. The All of Us Research Program is supported by the National Institutes of Health, Office of the Director.

## Ethics

This analysis used de-identified All of Us Research Program data (version 7) via the Researcher Workbench. The Howard University IRB determined the study *exempt* (Category 4). All activities complied with All of Us data use and publication policies.

## Data Availability

De-identified data are available via controlled access through the All of Us Researcher Workbench (https://www.researchallofus.org) for investigators at institutions with an active All of Us Data Use Agreement who complete required training.

## Patient/Participant Consent

Not applicable (de-identified data).

## Clinical Trial Registration

Not applicable (observational study).

## Prior Presentation

None.

## Supplementary Material

See Supplementary Tables S1–S10 for extended results and sensitivity analyses (provided as a separate PDF).

