## Supplementary Tables for "Medication-Stratified Analysis of LDL-C Equation Miscalibration in Diabetes: Evidence from the All of Us Research Program and a Medication-Agnostic Machine-Learning Correction"

for

#### Index of Supplementary Tables

1. **Table S1.** Medication Combination Matrix (sensitivity analysis)
2. **Table S2.** Additive Interaction (RERI) for insulin × GLP-1 on Equation Error
3. **Table S3.** Triglyceride Bands—Error Reduction vs. Friedewald and NRI
4. **Table S4.** Near-Threshold Misclassification and Abstention
5. **Table S5.** NRI@100 Breakdown by Subgroups
6. **Table S6.** No-equations ablation (equation features removed)
7. **Table S7.** Multivariable calibration with prediction × medication interactions
8. **Table S8.** Adding medication indicators at inference
9. **Table S9.** Permutation Feature Importance—Overall Cohort
10. **Table S10.** Permutation Feature Importance—GLP-1 Subset

### Supplementary Tables

*Note:* The following tables provide confirmatory evidence from a prespecified rerun of the identical pipeline (seed 123) on a resampled test split. They are referenced in the main text where noted.

Table 1: **Table S1.** Medication Combination Matrix (sensitivity analysis)

**Mean Absolute Error (MAE) by Medication Combination**

| **Combo** | $n$ | **ML** | **FW** | **NIH Eq2** | **MH** |
|---|---|---|---|---|---|
| I+G+S+ | 120 | 27.1 | 36.5 | 33.1 | — |
| I+G-S+ | 204 | 28.0 | 33.2 | 31.0 | — |
| I+G-S- | 22 | 20.5 | 25.3 | 22.4 | — |
| I-G+S+ | 24 | 27.5 | 44.1 | 39.1 | — |
| I-G+S- | 11 | 16.5 | 19.0 | 17.4 | — |
| I-G-S+ | 213 | 24.0 | 27.5 | 25.8 | — |
| I-G-S- | 97 | 18.0 | 19.3 | 18.3 | — |

**Calibration Slopes by Medication Combination**

| **Combo** | $n$ | **ML slope** | **FW slope** | **NIH Eq2 slope** | **MH slope** |
|---|---|---|---|---|---|
| I+G+S+ | 120 | 0.558 | 0.290 | 0.335 | — |
| I+G-S+ | 204 | 1.057 | 0.629 | 0.669 | — |
| I+G-S- | 22 | 0.522 | 0.339 | 0.382 | — |
| I-G+S+ | 24 | 0.854 | 0.330 | 0.368 | — |
| I-G+S- | 11 | 0.959 | 0.616 | 0.705 | — |
| I-G-S+ | 213 | 1.001 | 0.584 | 0.609 | — |
| I-G-S- | 97 | 0.969 | 0.615 | 0.650 | — |

MAE in mg/dL. "I"=insulin, "G"=GLP-1, "S"=statin; "+" indicates active exposure. "—" indicates Martin–Hopkins was not estimated due to sparse cell convergence issues (n<30 in these groups). Cells with n<30 are provided for descriptive context only and should not be interpreted causally.

Table 2: **Table S2.** Additive Interaction (RERI) for insulin × GLP-1 on Equation Error (sensitivity analysis)

| **Method** | **RERI** | **CI_lo** | **CI_hi** | *n* |
|---|---|---|---|---|
| Friedewald | −3.316 | −11.242 | 11.002 | 696 |
| Sampson (NIH) Equation 2 | −2.253 | −10.428 | 9.881 | 696 |

Wide confidence intervals spanning zero; descriptive evidence that combined insulin × GLP-1 exposure may compound equation bias.

Table 3: **Table S3.** Triglyceride Bands—Error Reduction vs. Friedewald and NRI (sensitivity analysis)

| **TG bin (mg/dL)** | *n* | **ML_MAE** | **FW_MAE** | **Δ% vs. FW** | **NRI@100 vs. FW** |
|---|---|---|---|---|---|
| (0, 150] | 469 | 23.975 | 25.491 | 5.948% | −0.011 |
| (150, 200] | 92 | 27.306 | 33.674 | 18.911% | 0.048 |
| (200, 300] | 84 | 26.377 | 37.445 | 29.558% | 0.106 |
| (300, 400] | 27 | 23.687 | 44.381 | 46.628% | 0.239 |
| (400, 2000] | 24 | 16.479 | 52.300 | 68.492% | 0.556 |

Table 4: **Table S4.** Near-Threshold Misclassification and Abstention (sensitivity analysis)

| **Threshold** | **near_n** | **mis_FW** | **mis_ML** | **abstained** |
|---|---|---|---|---|
| 70 | 93 | 48 | 45 | 38 |
| 100 | 67 | 32 | 29 | 21 |
| 130 | 48 | 19 | 19 | 15 |

Table 5: **Table S5.** NRI@100 Breakdown by Subgroups (sensitivity analysis)

| **Slice** | **Comparator** | **NRI** | **95% CI lo** | **95% CI hi** | $n$ |
|---|---|---|---|---|---|
| Overall | FW | 0.041 | −0.007 | 0.091 | 696 |
| Overall | NIH | 0.007 | −0.039 | 0.053 | 696 |
| TG≥200 | FW | **0.218** | **0.063** | **0.369** | 132 |
| TG≥200 | NIH | 0.076 | −0.065 | 0.208 | 132 |
| insulin=1 | FW | 0.079 | 0.003 | 0.150 | 380 |
| insulin=1 | NIH | 0.046 | −0.024 | 0.115 | 380 |
| glp1=1 | FW | 0.095 | −0.012 | 0.214 | 172 |
| glp1=1 | NIH | 0.045 | −0.046 | 0.148 | 172 |
| statin=1 | FW | 0.061 | 0.003 | 0.116 | 587 |
| statin=1 | NIH | 0.024 | −0.030 | 0.078 | 587 |
| AA=1 | FW | 0.026 | −0.076 | 0.129 | 160 |
| AA=1 | NIH | −0.027 | −0.114 | 0.064 | 160 |

Race subgroup rows are exploratory quality checks to ensure no gross calibration drift; they are not used to draw equity conclusions.

Table 6: **Table S6.** No-equations ablation (equation features removed)

| **Model** | **MAE** | **95% CI** | **Slope** | **95% CI** | **Intercept** | ΔMAE vs. base | ΔSlope vs. base |
|---|---|---|---|---|---|---|---|
| ML (no equations) | 24.66 | 23.02–26.20 | 0.89 | 0.78–0.99 | 11.30 | −0.03 | −0.02 |

Base (with equations): MAE 24.69 mg/dL, slope 0.91, intercept 8.82 mg/dL.

Table 7: **Table S7.** Multivariable calibration with prediction × medication interactions

| **Term** | **Coef** | **CI_lo** | **CI_hi** | **p** |
|---|---|---|---|---|
| pred | 0.9207 | 0.6228 | 1.2186 | $< 0.001$ |
| pred×insulin | −0.0621 | −0.3130 | 0.1888 | 0.6272 |
| pred×glp1 | −0.3033 | −0.5804 | −0.0263 | 0.0319 |
| pred×statin | 0.0645 | −0.2654 | 0.3945 | 0.7011 |

Most prediction × medication interactions were non-significant; a modest prediction × GLP-1 interaction (coef −0.30, $p = 0.032$) did not yield out-of-sample performance gains when medication indicators were added at inference.

Table 8: **Table S8.** Adding medication indicators at inference

| **Model** | **MAE** | **95% CI** | **Slope** | **95% CI** | **Intercept** | ΔMAE vs. base | ΔSlope vs. base |
|---|---|---|---|---|---|---|---|
| ML + med indicators | 24.75 | 23.06–26.29 | 0.86 | 0.76–0.97 | 13.81 | +0.06 | −0.05 |

Base (with equations, no med labels): MAE 24.69 mg/dL, slope 0.91, intercept 8.82 mg/dL.

Table 9: **Table S9.** Permutation Feature Importance—Overall Cohort

| **Feature** | **Importance** | **Std** |
|---|---|---|
| ldl_sampson_eqn | 0.3613 | 0.0352 |
| ldl_friedewald | 0.0146 | 0.0079 |
| is_male | 0.0113 | 0.0058 |
| age | 0.0110 | 0.0063 |
| is_obese | −0.0003 | 0.0003 |
| tg_hdl_ratio | −0.0024 | 0.0026 |
| tg_bmi | −0.0054 | 0.0045 |
| glucose | −0.0067 | 0.0053 |
| log_tg | −0.0068 | 0.0018 |
| tc_hdl_ratio | −0.0070 | 0.0036 |

Importance = mean decrease in accuracy; Std = standard deviation across 40 permutation repeats.

Table 10: **Table S10.** Permutation Feature Importance—GLP-1 receptor agonist subset

| **Feature** | **Importance** | **Std** |
|---|---|---|
| ldl_sampson_eqn | 0.3040 | 0.0744 |
| ldl_friedewald | 0.0361 | 0.0206 |
| is_male | 0.0317 | 0.0151 |
| age | 0.0243 | 0.0177 |
| hdl | 0.0075 | 0.0143 |
| glucose | 0.0070 | 0.0097 |
| bmi | 0.0058 | 0.0095 |
| tc_hdl_ratio | 0.0048 | 0.0086 |
| tg_hdl_ratio | 0.0014 | 0.0051 |

Importance = mean decrease in accuracy; Std = standard deviation across 40 permutation repeats.
